# Neurodiversity in the Paediatric Chronic Pain Clinic: An Audit

**DOI:** 10.64898/2026.06.02.26354725

**Authors:** Hadassah Buechner, Giorgos Themistokleous, Molly Orr, Eva Lawson, Eve Smart, Amy Donaghy, Ewan Wallace

## Abstract

**Objective:** To compare the characteristics, management and outcomes of neurodivergent (ND) children with neurotypical (NT) children attending a chronic pain clinic.

**Design:** An audit of all patients attending the clinic from 2010-2025 using electronic patient records.

**Setting:** A tertiary pain centre in Scotland.

**Patients:** 724 patients were included in the analysis, 193 (26%) were neurodivergent. Patients were included if they had a documented referral to the pain clinic and attendance to at least one clinic appointment. Patients were excluded if no pain clinic letter could be found on their records.

**Results:** There was a significant increase in the percentage of children with neurodiversity attending the chronic pain clinic compared to neurotypical children (*p* = 0.004) accounting for over a third of children last seen in the period of 2023-2025. ND children were most likely to present with musculoskeletal pain compared with NT children (*p* = 0.033) representing over half of all ND children’s presentations with pain. ND children were more likely to report being bedbound (18% ND, 13% NT, *p* = 0.0352) or needing a walking aid (40% ND, 25% NT, *p* = 0.000) due to chronic pain and had a higher number of referrals (ND median = 18.4, 1QR, NT median = 12.44, IQR10.28 *p* = 0.000). ND children were more likely to live in areas of deprivation (Cochran-Armitage test, Z –2.15, *p* = 0.0315).

**Conclusions:** Children with neurodiversity are overrepresented in the chronic pain clinic, and more often present to tertiary services with musculoskeletal pain. They are more likely to have multiple referrals, spend longer with the pain service and less likely to be discharged due to pain improvement. These findings highlight the need for focused strategies to address chronic pain in neurodivergent children. Services should consider how best to identify and support children with neurodiversity and chronic pain.

**Key Messages:** *What is already known on this topic:* While there has been research regarding the role of neurodiversity in pain perception, there are gaps in knowledge regarding the influence of neurodiversity on the development and persistence of chronic pain in children.

*What this study adds:* A growing proportion of neurodiverse children attended the pain clinic. Neurodiverse children presented with more severely impactful pain, they spent a longer duration of time within the pain clinic and were less likely to be discharged due to pain improvement.

*How this study might affect research, practice or policy:* Identifying neurodiverse children as a patient group with distinct requirements may prompt adaptations in chronic pain management practices. This audit provides an initial framework for subsequent research.

## Introduction

Chronic pain has been described as a “persisting or recurring pain lasting more than 3 months.” [1] Chronic pain may present without a clear underlying cause and can be a diagnosis itself. Chronic pain is common within paediatrics, with between 15-30% of children and adolescents experiencing daily pain. [2] Around 3% of these children experience disabling pain, and require referrals to tertiary services. [3] Children may experience loss of appetite, fatigue and sleep disturbances. [4] This can lead to feelings of frustration and hopelessness, which may contribute to developing anxiety or depression. A systematic review found that 35% of adolescence with chronic pain had anxiety disorders and 12% had depressive disorders, higher than the incidence in the general adolescent population. [5]

The impact of chronic pain extends beyond the individual to society, leading to pressure on social, economic and medical sectors. Analysis has shown that children between the ages of 5-18 with chronic pain are absent for 22% of school days [6]. Poor attendance at school has been linked to difficulties in social development, lower educational attainment and poorer prospects in employment. [7] Additionally, chronic pain in children can place emotional and financial strain on families, affecting parents’ work and wellbeing. [8]

The presentation of chronic pain is influenced by a range of biological, psychological and social factors. One factor that may contribute to severe or complex pain is neurodiversity. Neurodiverse individuals appear to be at a higher risk of experiencing chronic pain or having heightened pain sensitivity. [9] [10] Differences in sensory processing may amplify the perception of pain. Additionally, conditions such as depression, anxiety or sleep disturbances are more prevalent in neurodivergent populations which can contribute to the severity and persistence of pain. [11] Those with neurodiversity are also more likely to experience hypermobility, which has been associated with a higher incidence of musculoskeletal pain. [10]

Neurodivergence is used to describe a variation in the way that information is perceived, processed or expressed. [12] Neurodiversity was categorised in this audit as either autism spectrum disorder (ASD), attention deficit disorder (ADHD), awaiting assessment for ASD/ADHD, global developmental delay (GDD), learning difficulties, dyslexia, dyscalculia, dyspraxia, tic disorders or Down syndrome. The term neurodiversity is used in this audit because it is transdiagnostic and describes clinically relevant phenotypes. [13]

There is limited research exploring the link between chronic pain and neurodiversity. Understanding the relationship between neurodiversity and chronic pain is essential for addressing any incongruities in both recognition of pain and management within this group. By recognising that neurodivergent individuals may experience pain differently, services can address both the physical and psychological aspects of the pain, improving outcomes and quality of life. This exploratory audit aims to investigate the proportion of children with chronic pain who are neurodivergent (ND) and to compare their characteristics, care and outcomes to neurotypical (NT) children within a tertiary paediatric chronic pain service.

The aims of this audit are to:

- Examine differences in presentation, demographics, interventions received and clinical outcomes for neurodiverse children compared to neurotypical children.
- To identify areas of improvement which could support neurodiverse patients within chronic pain
- To provide an evidence base for future research into this area

## Methods

### Patients

Paediatric patients (aged 0-18) between 2010 and 2025 referred to the Paediatric Chronic Pain Service based in the Royal Hospital for Children Glasgow were consecutively audited up to 2025. Patients with no evidence of attending a chronic pain clinic were excluded (n = 64). Data gathered from 2023-2025 was prospective as the audit was ongoing during data collection.

### Data collection and reporting

Patient information via electronic patient records was collected using an 83-question standardised reporting survey. Patient information was gathered by the co-authors and was re-checked by co-authors to ensure fidelity. Pain was classified as per ICD-11. [1] Neurodiversity was categorised as autism spectrum disorder (ASD), attention deficit disorder (ADHD), awaiting assessment for ASD/ADHD, global developmental delay (GDD), learning difficulties, dyslexia, dyscalculia, dyspraxia, Tourette syndrome or Down syndrome. The survey covered questions on various aspects including sex, gender identity, Scottish Index of Multiple Deprivation (SIMD), ethnicity, pain history, co-morbidities, neurodivergence, hypermobility, referrals, management, family information, impact on activities of daily living, patient outcomes, and barriers to service usage. Adversity was classified as per the Felitti et al. [14] The manuscript was written following SQUIRE guidelines.

During the audit process the initial data was presented to the paediatric chronic pain medical and psychology team. Round table discussion between faculty and auditors sparked debate on how best to improve the service. Outputs included a drive to identify improvements for children with neurodiversity, and to increase the documentation of social factors via establishment of a clinic pro forma. Auditing continued for two years in total.

### Statistical analysis

Pearson’s chi-squared tests were performed for categorical variables. Welch’s t-tests or Mann-Whitney U test was used for continuous variables. A Cochran-Armitage linear effects model was used to assess trend for SIMD. Only basic statistics was used as this was an audit-based project. Statistical analyses were performed using R version 4.5.1, statistical significance set at p < 0.05.

## Results

### Incidence and Type of Neurodiversity

726 patients were included in the analysis. 26% of children seen in clinic with chronic pain were neurodivergent, n = 193. Autism spectrum disorder (ASD) was the most reported, with 51% of neurodiverse patients having a diagnosis of ASD. Learning difficulties were also prevalent, identified in 17% of patients. 10% of patients had a diagnosis of attention deficit hyperactivity disorder (ADHD), whilst 13% were awaiting assessment for ASD or ADHD. Few patients had documentation of global developmental delay, dyslexia, dyscalculia and dyspraxia. Patients within the “other” category had conditions such as Down’s syndrome and Tourette’s syndrome. Supplementary Table 1 illustrates the types of neurodivergence within this population.

### Age at first documentation of Pain

The median age of all patients when their chronic pain was first noted in medical records was 11. The mean age for neurodivergent children was 10.2 years and the mean age for neurotypical children was 10.9 years. This difference was statistically significant, *p* = 0.015.

### Sex and Gender

In the neurodivergent group, 61.1% of children were female and 37.8% were male. In the neurotypical group, 68.1% were female and 31.5% were male. Within both groups, female sex was more prevalent, this was significant (NT, *p* = 0.000 and ND, *p* = 0.001). There was no statistically significant difference in the percentage of male children between groups, although the ND group consisted of a higher percentage of males (p = 0.111). Data on gender identity was collected, with 4 children in the ND group identifying as non-binary, and no children in the NT group identifying as non-binary. “Unknown gender identity” was documented for 96 patients (13.3% of the whole sample). Demographics are summarised in supplementary table 2.

Patients were grouped into epochs depending on when they were last seen in clinic. The total number of patients in each epoch increased. Proportionally ND patient numbers grew more than NT patients, with the biggest increase seen between 2020-2022 and 2023-2025. By 2023-2025, ND patients accounted for more than a third of the children in the chronic pain clinic (table 1). The rate of increase in neurodiverse patients was significantly greater than the rate of increase of neurotypical patients from 2010 to 2025 (*p* = 0.004).

**Table 1:** Changes in Patient Cohort over the Years.

| Epochs | Neurodivergent | Percentage of patients | Neurotypical | Percentage of patients |
| --- | --- | --- | --- | --- |
| 2010-2016<br>N = 128 | 25 | 20% | 103 | 80% |
| 2017-2019<br>N = 138 | 30 | 22% | 108 | 78% |
| 2020-2022<br>N = 186 | 45 | 24% | 141 | 76% |
| 2023-2025<br>N = 274 | 93 | 34% | 181 | 66% |

### Type of Pain – ICD-11 pain types

In both groups secondary chronic pain was the most prevalent type of pain. Of ND children, 92/193 (47.7%) reported suffering from secondary chronic pain, compared with 252/533 (47.3%) NT children. Primary chronic pain was more common in the neurotypical group, accounting for 214/533 (40.2%) in NT, and 72/193, (38.3%) in ND. A small proportion of patients had evidence of primary and secondary pain diagnoses. There was no statistically significant difference in the type of pain based on neurotype (*p* = 0.5914).

### Location of Pain

Pain diagnosis by location were gathered. The five most common are reported in Figure 1 below. The most common complaint for all patients was musculoskeletal (MSK) pain, this includes both primary and secondary MSK pain diagnoses. The difference between the proportion of ND patients with MSK pain compared to NT patients with MSK pain was significant, *p =* 0.033. The differences between ND patients and NT patients in the other pain locations was not significant.

**Figure 1:**
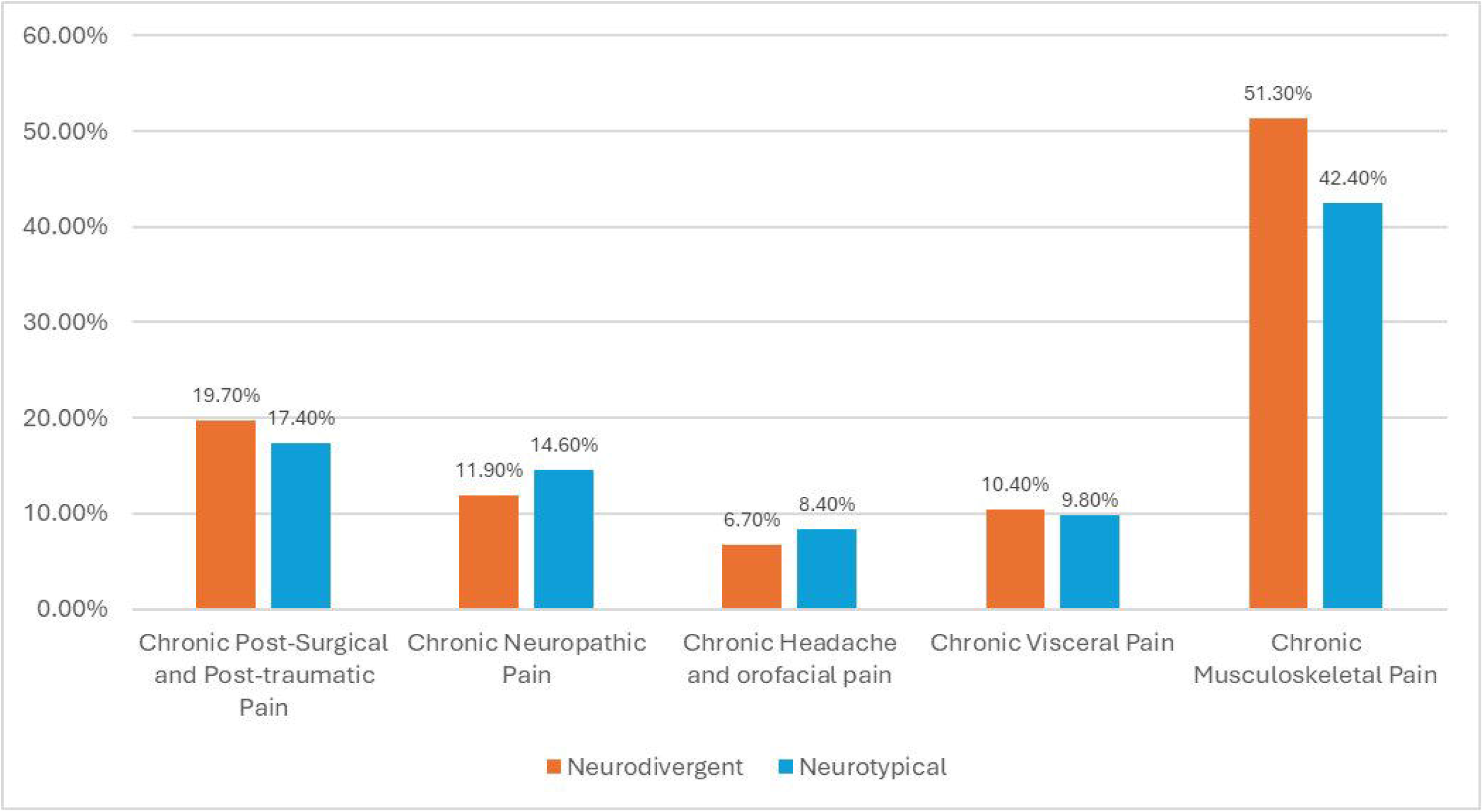
Location of Pain

**Figure 2:**
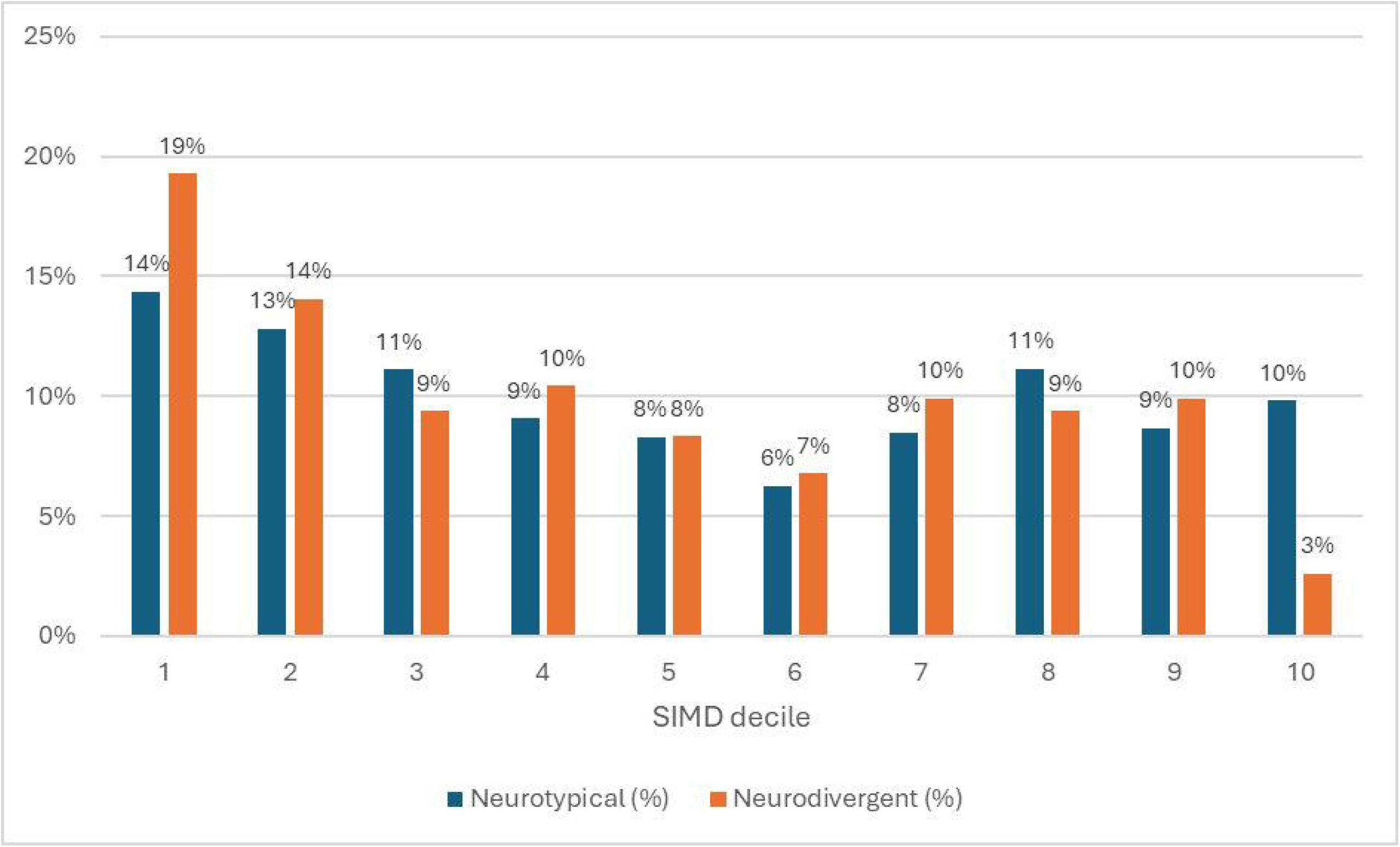
Distribution of ND and NT patients across SIMD deciles

### Scottish Indices of Multiple Deprivation (SIMD)

The SIMD decile ranking of the patients differed between the two groups. Those with neurodiversity were more likely to be living in a deprived areas than their neurotypical peers, (figure 3). Both the neurotypical and neurodiverse groups were most often from the 1st decile, the most deprived, at 14% and 19% respectively. The difference between the proportion of ND and NT children in the most deprived decile was not statistically significant (*p* = 0.148). Only 3% of neurodiverse patients were from the least deprived decile (10th), contrasting with 10% of the neurotypical group. The difference between the incidence of ND and NT patients in the least deprived decile was statistically significant (*<u>p</u>* = 0.0019). Logistic regression of the data suggests that fewer ND patients are found in affluent deciles, this trend neared significance (p = 0.0612). Cochran-Armitage test found a significant linear trend across the deciles, with ND patients more likely to be found in deprived deciles (*Z* – 2.15, p = 0.0315).

**Figure 3:**
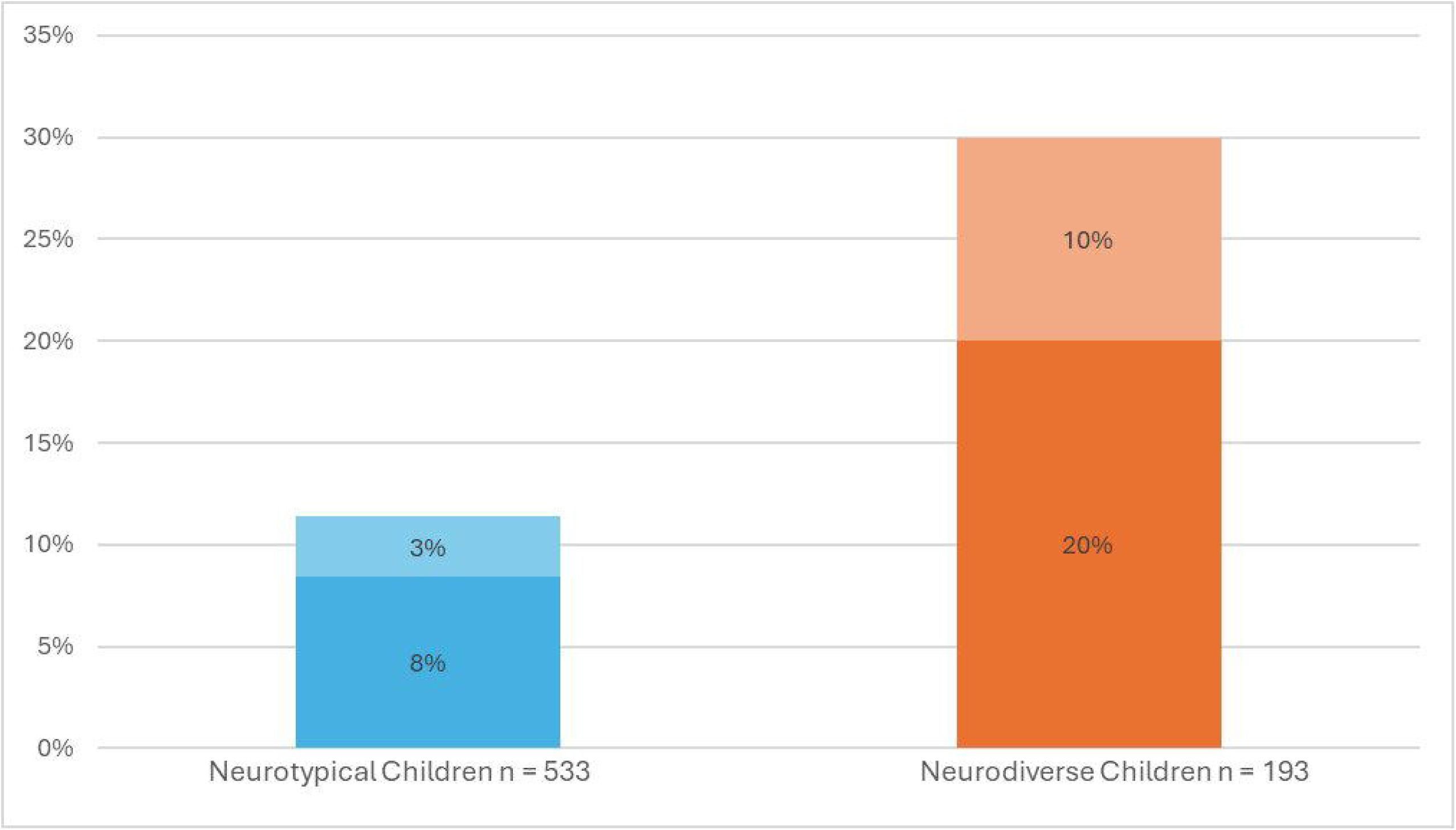
Reported Hypermobility in Children with Chronic Pain. Darker shade denotes female.

### Family History of Chronic Pain

Those diagnosed with a neurodiversity were more likely to have a parent with a chronic pain condition. 10.4% of the neurodiverse cohort reported parental chronic pain diagnoses, against 8.44% in the neurotypical cohort. By parent the most common co-occurring pain diagnosis was a biological mother (6.7% ND vs 6.6% NT), biological father (2.6% ND vs 1.5% NT), and both parents (1.0% ND vs 0.2% NT), this difference was significant (*p* = 0.02972). Regarding siblings, 4.66% of those with a neurodiversity reported having a sibling with a chronic pain condition, compared to 3.75% of the neurotypical cohort. This difference was not significant (*p* = 0.7345). See supplementary table 3 for a breakdown.

### Hypermobility

Hypermobility was higher in neurodiverse children (n = 58, 30.1%), compared to neurotypical children (n = 63, 11.8%), *p* = 0.000. Within both groups, females were more likely to be hypermobile. 20% of all ND children were hypermobile females compared with 10% hypermobile males. Of NT children, 8% were hypermobile females and 3% were hypermobile males, see figure 3.

### Adversity

Adverse childhood experiences include any kind of abuse, domestic violence or neglect. The data found 16.6% of neurodivergent children reported at least one adversity, compared with 15.0% in neurotypical children. This difference was not statistically significant (*p* = 0.8526).

### Functional Impact of Pain

The functional impact of pain was assessed based on the number reporting ever using walking aids or ever being bedbound due to pain. It was more common for patients in the neurodivergent group to have been bedbound (18.1% ND, vs 12.9% NT). ND children were more likely to report ever using a walking aid (39.9% ND vs 24.8% NT). The difference was statistically significant for use of a walking aid (*p* = 0.000) and for being bedbound (*p* = 0.0352).

### Referrals and Management

The total number of referrals the children had from any clinician was recorded and the difference in referral patterns between neurodivergent and neurotypical patients were analysed. Children in the ND group had a median of 15 referrals, IQR 17, mean of 18.39 referrals, σ = 15.72. The NT children had a median of 10 referrals, IQR 12, mean of 12.44, σ = 10.28. This difference was statistically significant at *p* = 0.000. The trend was similar for the number of GP referrals, a subset of the total referrals. ND children had a median of 4 GP referrals, IQR 8, mean of 6.76, σ = 7.33. NT children had a median 4, IQR 7, mean of 5.34 GP referrals, σ = 5.59, (*p* = 0.122).

### Referrals per Specialty

Referrals to almost all specialties were higher in ND children. Neurodivergent children were more often referred to General Paediatrics, at 52.9% of all ND children, compared to 45.2% of NT group, (*p* = 0.083). Similarly, Physiotherapy referrals were more common in the ND group (52.3%) compared with the NT group (43.1%), (p = 0.035). Psychiatry also showed a disparity with 37.3% compared to 25.89%, (p = 0.004). Among other specialist services, ND children were more frequently referred across multiple areas, the significance of the differences are summarised in table 2, see supplementary figure 2 for a visual overview.

**Table 2:** Referrals to other Specialties.

| Specialty | Percentage Neurodiverse referred (%) | Percentage Neurotypical referred (%) | $p$ - value |
| --- | --- | --- | --- |
| General Paediatrics | 52.85 | 45.22 | 0.083 |
| Physiotherapy | 69.3 | 53.5 | 0.035* |
| Psychiatry | 37.31 | 25.89 | 0.004* |
| Rheumatology | 38.5 | 28.3 | 0.008* |
| Neurology | 36.27 | 20.64 | 0.000* |
| Orthopaedics | 51.3 | 46.9 | 0.336 |
| Ophthalmology | 22.28 | 18.39 | 0.287 |
| Dietetics | 21.24 | 6 | 0.000* |
| Cardiology | 18.13 | 12.95 | 0.100 |
| Orthotics | 23.4 | 10.3 | 0.030* |
| Gastroenterology | 24.35 | 16.32 | 0.019* |
| Dentistry | 22.8 | 12.2 | 0.001* |
| Endocrinology | 18.13 | 11.63 | 0.031* |
| Occupational Therapy | 12.95 | 6.38 | 0.007* |
| Audiology | 9.84 | 5.63 | 0.067 |
| SALT | 3.63 | 1.69 | 0.149 |
| Genetics | 11.4 | 5.82 | 0.017* |
| Dermatology | 26.6 | 28.5 | 0.640 |
| Oncology | 2.1 | 2.4 | 0.780 |
| Podiatry | 17.7 | 16.7 | 0.839 |

### Medications

Patterns of medication use were similar between neurotypes. The most prescribed agents in both groups were NSAIDs (71.9% ND vs. 70.7% NT) and paracetamol (71.4% ND vs. 71.7% NT). Likewise, use of gabapentin was also similar between groups (33.3% ND vs. 32.1% NT). In contrast, melatonin was prescribed more frequently in ND children (22.3%) compared with NT children (7.1%), *p* = 0.000. Smaller differences were seen in co-codamol (21.9% ND vs. 19.5% NT) and amitriptyline (34.9% ND vs. 38.3% NT), although rates were broadly comparable. Opioid medication use (excluding co-codamol) was slightly lower in ND children (17.7%) compared with NT children (20.8%).

### Other management

The prescription of TENS machines (35.9% ND vs. 35.3% NT, *p* = 0.847), and psychological therapies (32.8% ND vs. 37.5% NT, *p* = 0.258) showed minimal difference between groups.

### Years Spent with the Chronic Pain Service

The distribution of years spent with the paediatric pain service before being discharged differed between neurotypical and neurodivergent children. Neurodivergent children spent a longer time with the paediatric pain service, with a median of 3 years, compared to 2 years in neurotypical children, (figure 6). A greater proportion of neurodivergent children continued to access the service for up to 7 years (7.4%), 8 years (4.4%), and even 9 years (0.7%). There was no statistically significant difference in the mean number of years children spent in chronic pain service (*p* = 0.2672).

### Outcomes

Patient outcomes were analysed. These outcomes included: still being in the service, being discharged, or no documentation. ND children were more likely to still be in service, with 20.3% of all ND children still in service, compared with 15.95% of NT children. This difference was not statistically significant (*p* = 0.18).

Reasons for patient discharged were analysed. 427 NT children (80.4%) and 143 ND children (74.1%) were included in the discharge analysis. Reasons for discharge included: pain improvement, patient non-engagement, the service being not effective, transferring to adults, referred to alternative service for pain or “other.” NT children were more likely to be discharged due to pain improvement (38.2% of discharged NT children vs 22.8% of discharged ND children). Discharge due to the service being no longer effective was similar between groups at 5.7% and 5.8% respectively. NT children were more likely to be discharged due to non-engagement with services (18.9% vs 14%). ND children were slightly more likely to be transferred to adult services (12.9% vs 9.8%). Discharge due to referral to alternative services was similar between groups. The “other” category included reasons such as not documented, patient being deceased, or patient moving to another health board. ND children more often had “other” reported in their notes, accounting for 38.6%, compared with 20.7%. The difference between groups for discharge reasons was statistically significant (p = 0.000), see figure 7.

### Barriers to Engaging with the Pain Service

Barriers to engagement with the paediatric pain service were more common among neurodivergent children. 14.51% (n = 28) of neurodivergent children had documented barriers compared to only 8.82% (n = 47) of neurotypical children, (*p* = 0.037). The nature of the barriers was similar between groups. The most frequent barrier reported was patients not being able to attend appointments. Some patients declined treatment or expressed distrust towards the healthcare system, which often led to discharge. Other barriers related to difficulties engaging with the service, for example due to mental health issues.

## Discussion

This audit identified children in the chronic pain service who had evidence of neurodiversity. The proportion of neurodivergent children was high, at 26% overall, reaching 34% of children seen in the last three years (table 1). Around 16% of Scottish children are thought to be neurodivergent, with 2.6% having a diagnosis of autism. [15] The higher rates of neurodiversity in the pain clinic are consistent with existing literature highlighting the increased vulnerability of neurodiverse people. [16], [17], [18], [19]

### The Presentation of Pain

Neurodivergent children in the service experienced more chronic MSK pain than neurotypical children, accounting for more than half of chronic pain conditions in the ND group (figure 1). Pain has been shown to be influenced by factors such as emotional dysregulation, sensory sensitivity and hypermobility in children with autism spectrum disorder. [9] The increased incidence of hypermobility in the audit’s neurodivergent patients (figure 3) could provide an explanation as to why there were higher rates of chronic MSK pain found. An association between hypermobility, pain and neurodiversity is consistent with existing literature. [10], [20] It has been proposed that the increased incidence of hypermobility in neurodivergent children is due to the pleiotropic nature of genetic variants associated with connective tissues and molecules expressed in the central nervous system. [10], [21] There is evidence that children of parents with chronic pain conditions are more likely to experience a chronic pain condition themselves. [22] Research into parental factors has suggested that behaviours within the family influences the pain experience. [23] This audit demonstrated that a family history of chronic pain conditions was more common in ND children, which is likely due to an interaction of innate and social factors. [24], [25]

### The Impact of Pain

The functional impact of pain was assessed by reported use of walking aids or being bedbound due to pain. Neurodivergent children were significantly more likely to report being bedbound and using a walking aid than neurotypical children (figure 4). Walking aids can be used for a variety of conditions, and the use of walkers and wheelchairs may allow for children to participate in more physical activity. There is a gap in the evidence for long-term effects of walking aids on overall health, compared to approaches to manage pain without aids. [26], [27]

**Figure 4:**
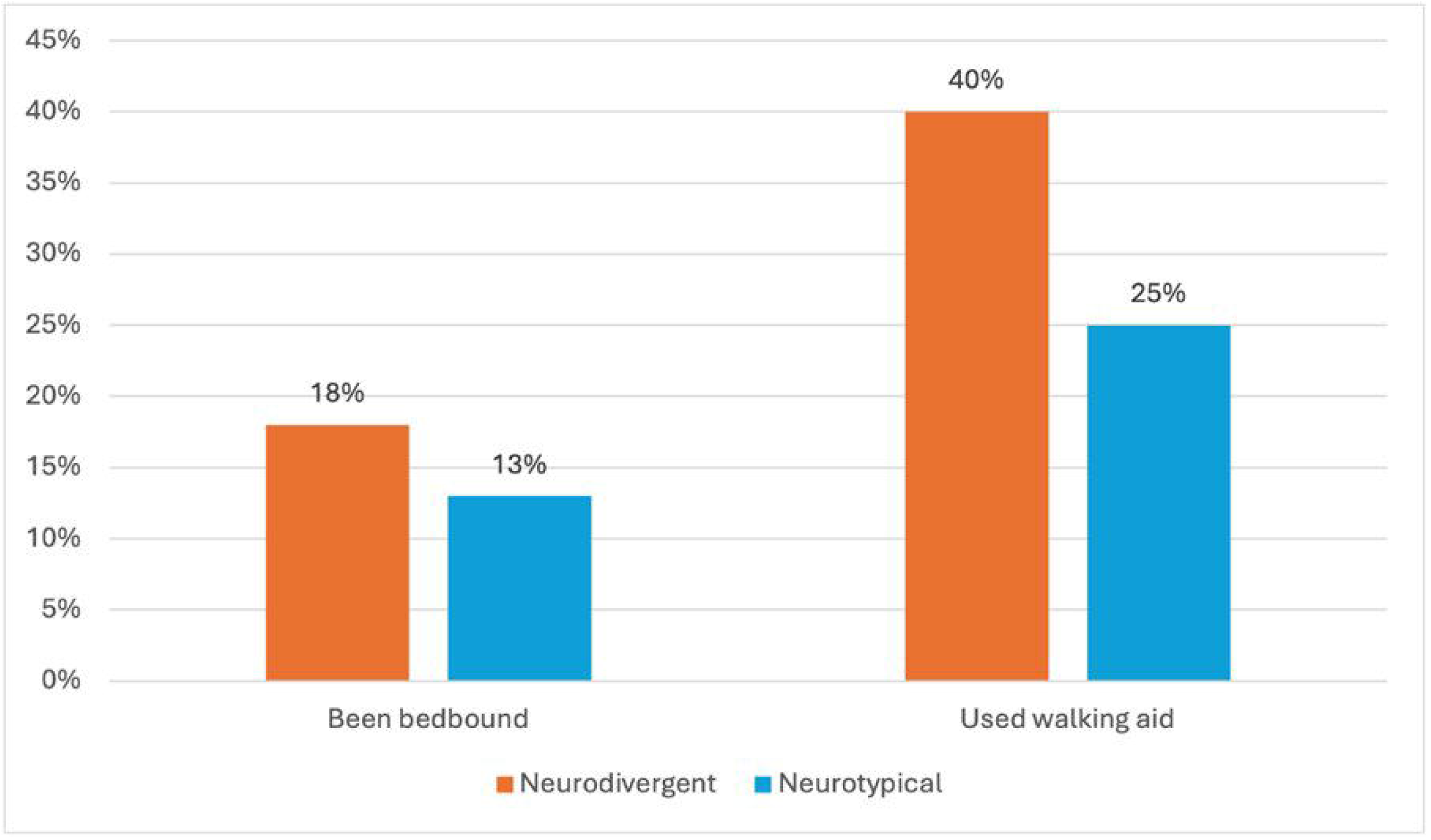
The Functional Impact of Chronic Pain

### Referrals

This audit identified significant differences in the pattern of referrals observed between neurodivergent and neurotypical children. Overall neurodivergent children were consistently referred to a higher number and broader range of specialities, such as general paediatrics, psychiatry, physiotherapy, and others (figure 5). The range of referrals reflects the complex clinical needs in neurodivergent children, who may face physical, psychological, and neurodevelopmental concerns. Neurodiversity may more often co-occur with epilepsy, gastrointestinal problems, mental illness, and different eating behaviours, accounting for the higher rates of referrals to neurology, gastroenterology and dietetics. [28], [29], [30]

**Figure 5:**
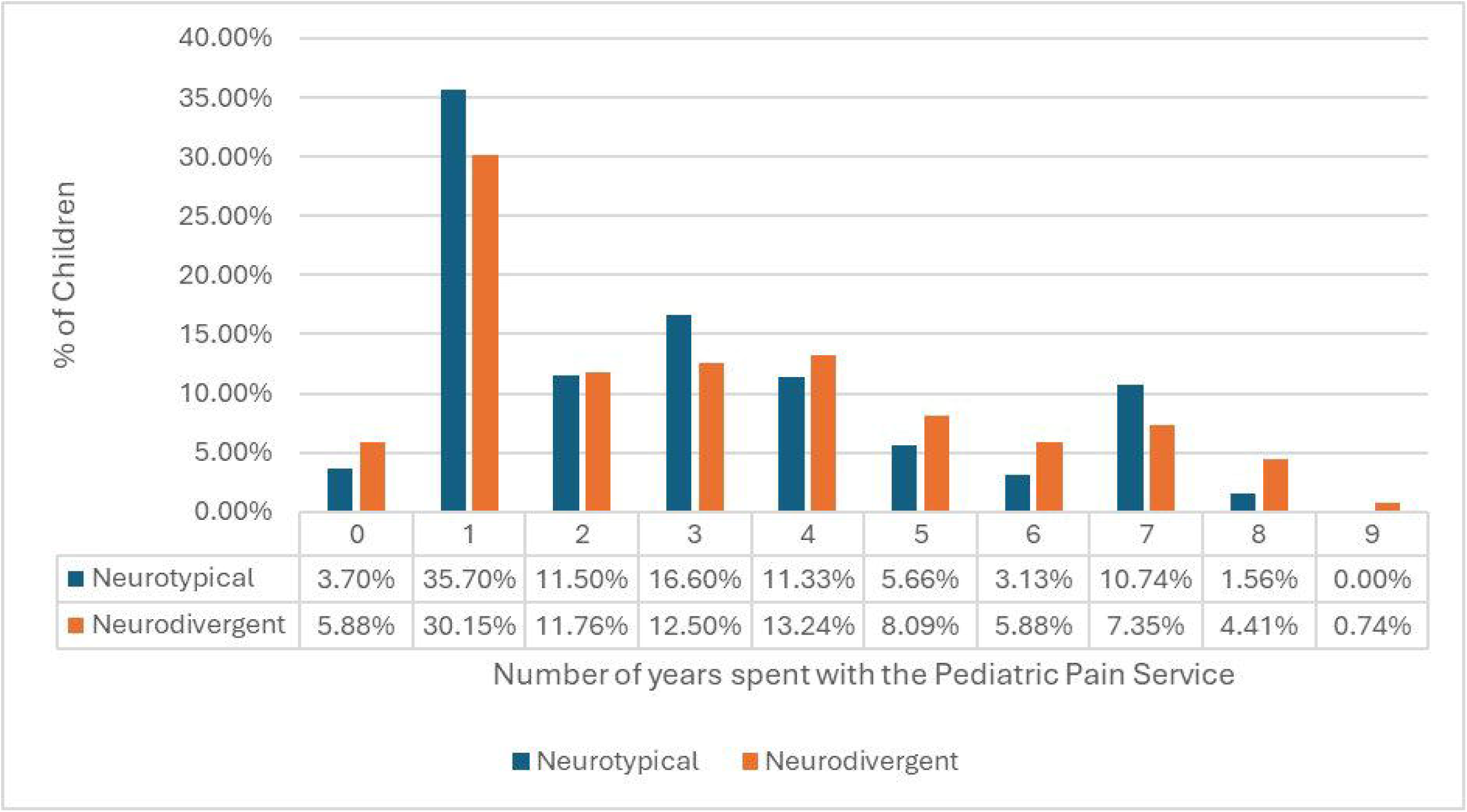
Length of time spent with the paediatric pain service

**Figure 6:**
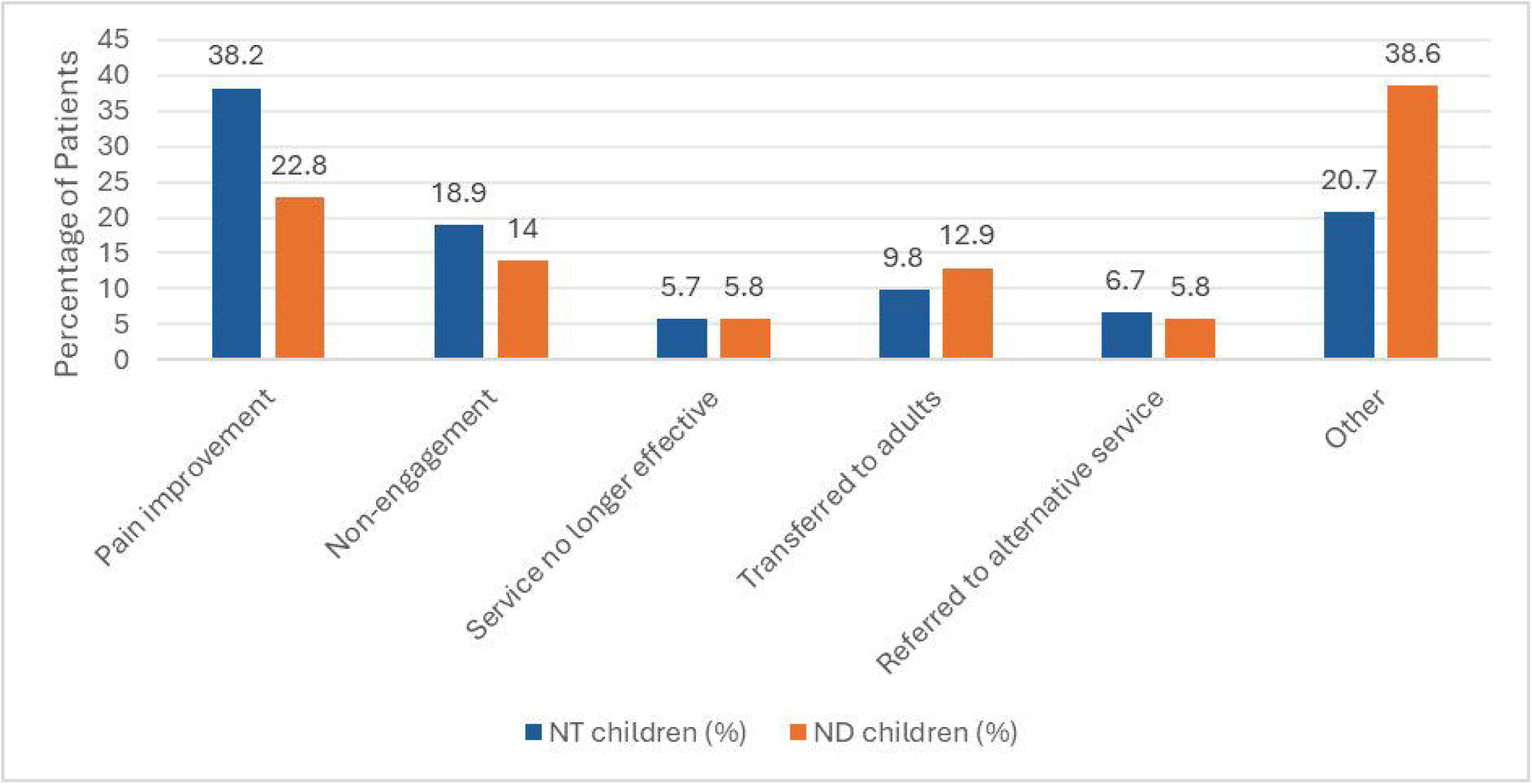
Reason for Discharge by Neurotype

The high referral count raises the concern of fragmented care, disjointed management and longer waits, challenges identified in Scottish adult pain services. [31] The results suggest the family of ND children are attending multiple appointments. A more integrated approach, such as MDT clinics and neurodiversity-informed pathways, could support coordinated management for neurodivergent individuals and their families.

### Medical and Alternative Management

The management between the patient groups was similar, with near-identical use of paracetamol, NSAIDs, gabapentin, opioids, TENS machines and psychological therapies. This shows a universal management pathway for chronic pain regardless of neurotype. The key difference in the pharmacological prescription for chronic pain was melatonin, which was three times more likely for to be given to neurodivergent children than their neurotypical peers. This difference reflects evidence of the high prevalence of sleep difficulties in neurodivergent people. [32] Melatonin is currently primarily indicated in the UK for children with neurodiversity where behavioural approaches have failed to improve sleep. [33] Poor sleep has been shown to exacerbate pain sensitivity and reduce pain tolerance, so melatonin may aid pain management. [34] It should be noted that behavioural approaches to sleep management should be prioritised initially. [35]

### Psychosocial Factors

This study found that neurodivergent children were more likely to come from socioeconomically deprived backgrounds, as measured by the Scottish Index of Multiple Deprivation (SIMD). Almost one-fifth of neurodivergent children in this study were from the most deprived decile, compared to 14% of neurotypical children. 3% of neurodivergent children were from the most affluent decile, compared to 10% of their neurotypical peers. Socioeconomic deprivation is a well-recognised determinant of health. The audit results support evidence linking financial hardship, chronic pain and neurodivergence. [36], [37]

Adversity was more frequently documented among neurodivergent children in this study. Extreme financial hardship could also be considered as an environmental adversity, [38] although it was not classified as one in this audit. Exposure to adverse childhood experiences has been linked to a dysregulated stress response, heightened pain perception and an increased likelihood of developing chronic pain. [39] The adversity noted to be more common in neurodivergent children may exacerbate both the severity and persistence of pain and could lead to poorer outcomes.

### Outcomes

Neurodivergent children were significantly less likely to be discharged from clinic due to pain improvement than neurotypical children (figure 7). They were less likely to be discharged for any reason, which aligns with the data showing ND children spending an average of a year longer in the service than NT peers (figure 6). An initially more severe, or severely impactful, pain presentation in ND children may explain why fewer children are discharged due to pain improvement.

Pain and neurodiversity interact in a way which leads to greater complexity. Children were noted to have more barriers to service engagement if they were neurodivergent. Communication challenges and atypical pain expression associated with neurodiversity could lead to these barriers. [40], [41] Higher appointment burden may increase the chance of appointment non-attendance. The increased likelihood of neurodivergent children in the pain clinic coming from areas of socioeconomic deprivation further exacerbates the difficulties faced by these children with regards to their wellbeing, pain management or pain remission.

### Strengths and Limitations

This audit is one of the first to interrogate the differences in pain presentation and service provision between neurodivergent and neurotypical children. A comprehensive analysis of all documented evidence ensured medical and psychosocial factors, where known, were included. This was an audit of a tertiary service in Scotland and may not be translatable to other centres. Additionally, few clinicians documented enquiry about social factors, such as adverse childhood experiences. Therefore, the recording of no adversity present, or of insufficient documentation, was at the discretion of the auditors, based on the contents of the clinical letters. It is likely that the number of adversities reported is an underestimate.

### Conclusion: Implications for Research and Recommendations

This audit highlights that neurodivergent children are disproportionately represented in the chronic pain clinic. The differences between neurodivergent and neurotypical children suggest that children with neurodiversity and chronic pain should be considered as a unique group of patients, accounting for over a third of children seen in the last three years. Services should consider pro-actively triaging neurodiverse children to MDT clinics and should consider whether any adaptations to support children with neurodiversity may be beneficial. Investment in additional education or training on neurodiversity for staff working with children who have chronic pain could be worthwhile. Specific resources aimed at children with chronic pain and neurodiversity, could be provided. Based on the incidence of neurodiversity in our audit, clinicians in tertiary pain services should remain open to the possibility that their patient may be neurodiverse, even if not diagnosed. Finally, dedicated audits of the management and outcomes of children with neurodiversity and chronic pain may provide areas to focus on in future quality improvement work.

## PPI statement

No members of the public or patients were involved due to the clinical nature of this audit.

## Data Availability

All data produced in the present study are available upon reasonable request to the authors

## Acknowledgements

The co-authors would like to thank Dr Shreena Unadkat and Miss Ginevra Bubani for their support during the audit.

## Ethical considerations

The audit was conducted in accordance with ethical principles and data protection practices. All data was anonymised prior to analysis to ensure participant confidentiality. As this audit utilised existing clinical data, informed consent was not required.

