## Supplementary tables for "Neurodiversity in the Paediatric Chronic Pain Clinic: An Audit"

| **Type of Neurodivergence** | **Number** | **Percentage** |
| --- | --- | --- |
| ASD | 98 | 51% |
| ADHD | 19 | 10% |
| Awaiting assessment for ASD/ADHD | 25 | 13% |
| GDD | 11 | 6% |
| Dyslexia | 20 | 10% |
| Dyscalculia | 4 | 2% |
| Dyspraxia | 5 | 2% |
| Learning Difficulties | 34 | 17% |
| Others | 38 | 19% |

**Supplementary Table 1:** Types of Neurodiversity in Children with Chronic Pain

| Demographic | Neurotypical n = 533 | Neurodivergent n = 193 | *p*-value |
| --- | --- | --- | --- |
| Mean Age at first pain documentation (years) | 10.9 | 10.2 | 0.015* |
| Biological Sex | | |  |
| Male | 168 (31.5%) | 73 (37.8%) | 0.111 |
| Female | 363 (68.1%) | 118 (61.1%) |  |
| Gender Identity | | |  |
| Male | 139 (26.1%) | 71 (36.8%) | 0.000* |
| Female | 314 (58.9%) | 96 (49.7%) |  |
| Non-Binary | 0 (0%) | 4 (2.1%) |  |
| Unknown | 78 (15%) | 22 (11.4%) |  |
| Ethnicity | | |  |
| White British | 380 (71.7%) | 141 (73.1%) | 0.581 |
| White Irish | 2 (0.4%) | 0 (0%) |  |
| White Other | 20 (3.8%) | 14 (7.3%) |  |
| White and Black African | 0 (0%) | 1 (0.5%) |  |
| Other mixed ethnicity | 6 (1.1%) | 1 (0.5%) |  |
| Indian | 2 (0.4%) | 0 (0%) |  |
| Pakistani | 13 (2.5%) | 5 (2.6%) |  |
| Chinese | 1 (0.2%) | 0 (0%) |  |
| Other Asian background | 2 (0.4%) | 1 (0.5%) |  |
| Black African | 4 (0.8%) | 0 (0%) |  |
| Other Black/African/Caribbean background | 2 (0.4%) | 1 (0.5%) |  |
| Arab | 1 (0.2%) | 0 (0%) |  |
| Any other | 3 (0.6%) | 1 (0.5%) |  |
| Unknown | 94 (17.7%) | 29 (15.0%) |  |

**Supplementary table 2:** Demographics

| Parental history of chronic pain condition | Neurotypical cohort (count, %)  n = 533 | Neurodiverse cohort (count, %)  n = 193 | *p*-value |
| --- | --- | --- | --- |
| Biological Mother | 35, 6.6% | 13, 6.7% | 0.03* |
| Biological Father | 8, 1.5% | 5, 2.6% |  |
| Both | 1, 0.20% | 2, 1.0% |  |
| Other | 1, 0.20% | 0, 0.00% |  |
| None/not documented | 394, 73.9% | 121, 62.7% |  |
| Sibling history of chronic pain condition | Neurotypical cohort (count, %)  n = 533 | Neurodiverse cohort (count, %)  n = 193 | p-value |
| Yes | 20, 3.75% | 9, 4.66% | 0.734 |
| None/not documented | 513, 96.25% | 184, 95.34% |  |

**Supplementary table 3:** Family History of Chronic Pain
