## Supplementary figures and images for "Neurodiversity in the Paediatric Chronic Pain Clinic: An Audit"

### Supplementary figure 1

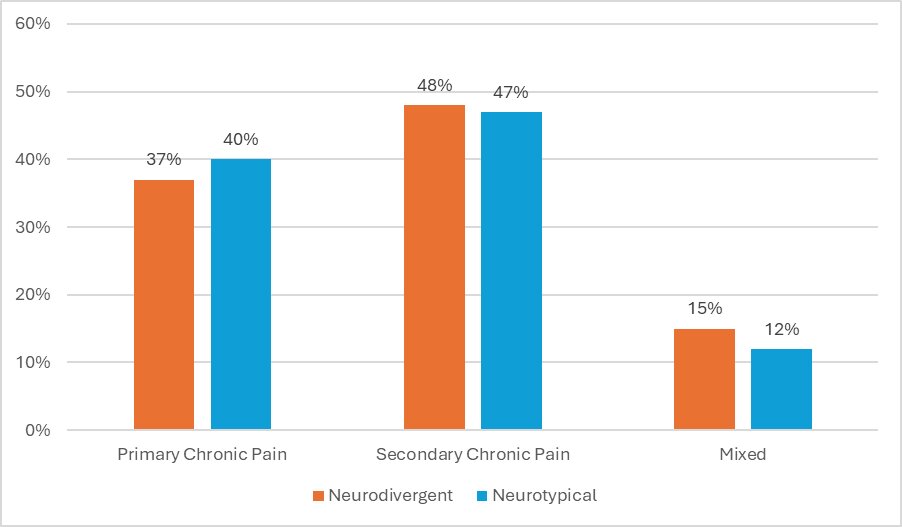

### Supplementary figure 2

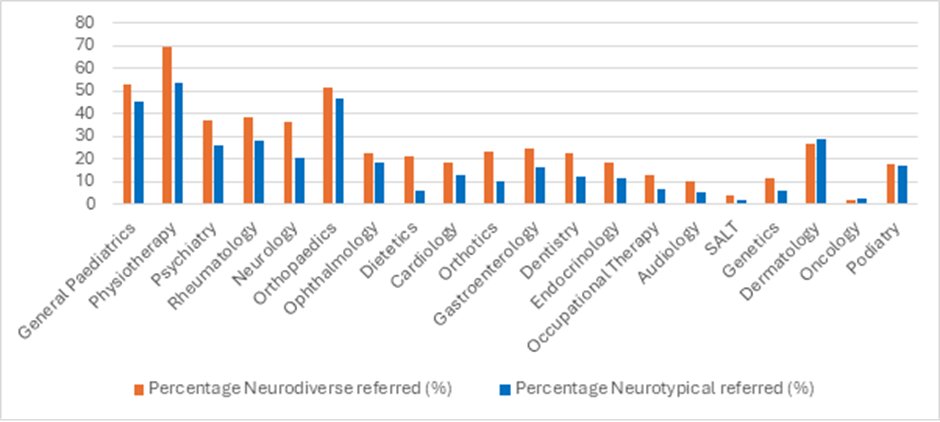
